# Naked-Eye Detection of LAMP-Produced Nucleic Acids in Saliva using Chitosan-capped AuNPs in a Single-Tube Assay

**DOI:** 10.1101/2023.06.09.23291198

**Authors:** Stylianos Grammatikos, Ioannis Svoliantopoulos, Electra Gizeli

## Abstract

Loop-mediated isothermal amplification (LAMP) is a low-technology molecular assay highly adaptable to point-of-care (POC) applications. However, achieving sensitive naked-eye detection of the amplified target in a crude sample is challenging. Herein, we report a simple, yet highly efficient and sensitive methodology for the colorimetric visualization of a single target copy in saliva, using chitosan-capped gold nanoparticles (Chit-AuNPs) synthesized via a green chemistry approach. The presence or absence of free Chit in the Chit-AuNPs solution was shown to affect LAMP colorimetric detection oppositely: the observed stabilization in the negative samples and aggregation in the positive samples in the presence of free Chit was reversed in the case of neat Chit-AuNPs. The mechanism of the two assays was investigated and attributed to electrostatic and depletion effects exerted between the Chit-AuNPs, free Chit and the solution components. The developed contamination-free, one-tube assay successfully amplified and detected down to 1-5 cfu of *Salmonella* and 10 copies of SARS-CoV-2 per reaction (25 μL) in the presence of 20% saliva, making the method suitable for POC applications. Compared to the commonly used pH sensitive dyes, Chit-AuNPs are shown to have an enhanced sensitivity toward the naked-eye colorimetric observation owing to the direct detection of DNA amplicons. Thus, this is a simple, highly sensitive, fast and versatile naked-eye detection methodology that could be coupled to any LAMP or RT-LAMP assay, avoiding the need of using complicated sample pretreatments and/or AuNPs long and laborious functionalization processes.

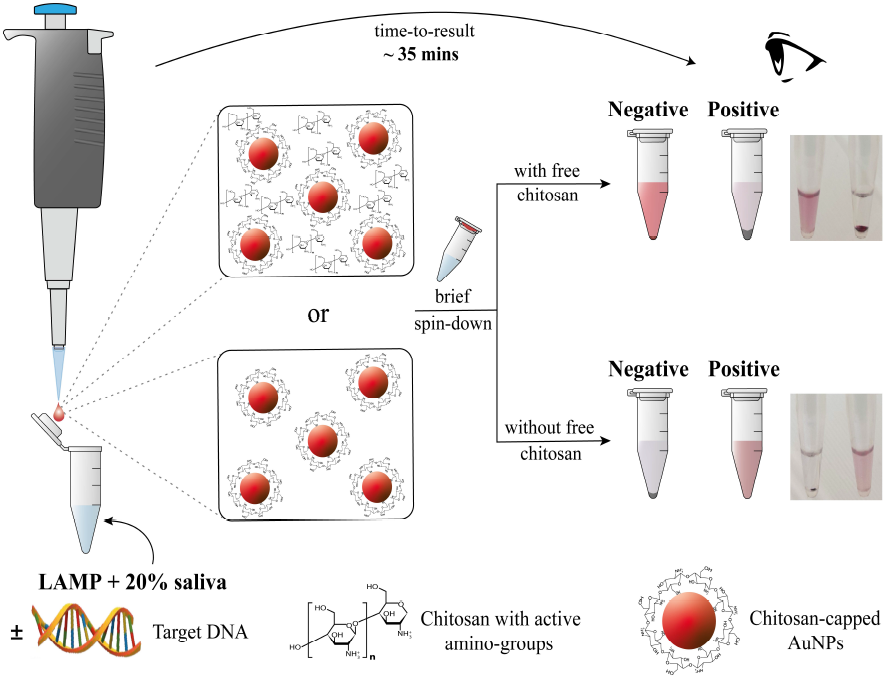

## Introduction

Loop mediated isothermal amplification (LAMP) has become a powerful alternative to polymerase chain reaction (PCR) for pathogen detection in clinical specimens^1,2^ and food matrices.^3,4^ Owing to the ability of Bst DNA polymerase to display a high strand displacement activity at a constant temperature (60-65 °C), isothermal LAMP has the potential to revolutionize molecular biology using simpler instrumentation, as well as faster and more efficient assays than PCR, even for crude samples.^5^ Consequently, LAMP has been adopted as a low-technology molecular analysis tool for resource-limiting areas and point-of-care (POC) applications.^6^ Achieving efficient detection at the POC, though, depends largely on the method selected for amplicons detection.

Naked-eye monitoring is the most promising method for inexpensive and simple diagnostics, with colorimetric detection being a prevalent approach for POC applications.^7^ The most common way to induce a nucleic acid-dependent colorimetric change is to use pH sensitive dyes by directly integrating them into a LAMP reaction;^8,9^ this is key in order to reduce the risk of contamination and provides a simple color-discrimination between the positive and negative samples. The potential drawbacks of using pH-sensitive dyes are the inability of the assay to generate sufficient pH variation, as reported for some targets and sample-types,^10^ and the need for a weakly buffered solution that restricts their use in the LAMP reaction.^11^ An alternative to pH-sensitive dyes is metallic nanoparticles (NPs), which can exhibit a size- and/or shape-dependent surface plasmon resonance (SPR) in the visible spectrum.^12^ Gold NPs (AuNPs), used extensively due to their strong quantum size effect and high stability, are capable of generating naked-eye visible color changes (e.g., from red to blue/purple, red to pink, and contrariwise)^13,14^ as a result of the NPs stabilization (dispersed) or destabilization (aggregated) induced by the presence or absence (or *vice versa*) of the target. There are two main approaches for AuNPs-based visual detection of amplicons, the target-specific and target-independent method.^7^ The former involves AuNPs functionalized with oligonucleotide (OG) probes where the state of the AuNPs relies on the complementarity of the probe with the target nucleic acid. The target-specific method has been used in combination with LAMP assays to detect different pathogens. The reported detection limits of the target-specific methods have been within the range of 10 to 200 copies per reaction;^15,16^ however, this was achieved upon DNA extraction, which is still a lab-based methodology. Recently, AuNPs-based target-specific assays have been developed in combination with the trans-cleavage activity of CRISPR/Cas systems, providing distinct colorimetric differences between the positive and negative samples, either by reverting or by promoting AuNPs aggregation.^17–19^ Notable drawbacks of the target-specific naked-eye colorimetric assays are the need for laborious and time-consuming protocols, including the need for the AuNPs surface-functionalization and/or multiple sample manipulation steps, e.g., mixing the amplified product with the AuNP probes, incubation and later reopening for salt addition, resulting in a contamination risk.

The target-independent detection method primarily relies on the stabilization or destabilization of AuNPs, owing to their electrostatic interactions. When AuNPs were coated with 11-mercaptoundecanoic acid (MUA), they aggregated in the presence of magnesium ions (Mg^2+^) in a negative sample while they were stabilized when magnesium pyrophosphate (Mg_2_P_2_O_7_), one of LAMP by-products, was formed in a positive sample (red solution).^20^ Furthermore, when AuNPs were co-functionalized with MUA and polyethylene-glycol (PEG), they were stable in the presence of Mg^2+^ in a negative sample due to steric hindrance (red solution), while they aggregated in the presence of Mg_2_P_2_O_7_ in a positive sample (red precipitate).^21^ The aforementioned assays have been demonstrated for the purified samples of extracted DNA with reported detection limits of 200 and 500 copies per reaction, respectively. An advantage of these assays is the ability to incorporate the AuNPs in the LAMP solution, although sonication is occasionally necessary to prevent aggregation; disadvantages include the dependency on the Mg_2_P_2_O_7_ by-product and the requirement of AuNPs surface functionalization. Another target-independent approach relies on biotinylated primers combined with streptavidin-coated AuNPs.^22,23^ This approach has been employed to generate a color change between the positive and negative samples; however, such assays present increased overall complexity due to multiple preparation steps.

Cationic polymers provide another potential means for the target-independent detection of amplified DNA via electrostatic interactions. An early study^24^ demonstrated this concept when positively charged polyethylenimine (PEI) was added to the LAMP reaction with a fluorescent-labeled OG probe, allowing visualization down to 0.2 μg of lambda DNA target in a purified sample. The main drawbacks of the method are increased contamination risk because of adding PEI after LAMP and the requirement of a UV illuminator and fluorescent probes for optical detection. In another study,^25^ chitosan (Chit) polysaccharide together with AuNPs were used for the naked-eye detection of mycobacterium tuberculosis using as starting material ∼30-400 μg/mL of whole DNA extracted from sputum. However, this assay was demonstrated in combination with PCR, a lab-based method using purified DNA; thus, it is unsuitable for POC applications.

Chitosan, a polysaccharide derivative of chitin sourced from the seafood industry, is a promising polymer because its cationic form (pH< pKa of ∼6.5)^26^ can conjugate DNA electrostatically, forming a Chit-DNA complex.^27,28^ The use of Chit-conjugated AuNPs has been reported in various applications, such as biosensing,^29^ drug delivery^30^ and tumor targeting,^31^ among others. Furthermore, Chit-coated magnetic NPs have been extensively used to extract DNA in acidic environments in which their charge is positive, and then release it in more basic environments, in which their charge gets neutralized.^32,33^ A main advantage of Chit-capped AuNPs is the ability to synthesize them using a green synthetic procedure. Chitosan is a non-toxic, eco-friendly, biosafe and biodegradable material.^34^ Moreover, it has been demonstrated to act both as a reducing and stabilizing agent, in a simple AuNPs synthesis procedure based on chemical reduction in aqueous environments;^35^ this method does not require toxic solvents or extra reducing agents. Although Chit appears promising for DNA-binding combined with aggregation/stabilization and colorimetric detection, its compatibility with LAMP has been low.^36^ To the best of our knowledge, Chit-capped AuNPs visual observation of LAMP products has not been reported yet. Overall, despite recent advances, the development of simple and economic colorimetric assays for naked-eye detection with high sensitivity still remains a challenge, i.e., detecting a few copies of the target in a crude sample with a low contamination risk (single-tube assay).

Herein, we report the development of a simple, fast and cost-effective method for naked-eye colorimetric detection of amplified nucleic acids produced via LAMP, using synthesized Chit-AuNPs and *Salmonella* enterica serovar Typhimurium as the selected target for demonstrating the proof of principle. Using a facile, rapid green synthesis protocol, positively charged Chit-AuNPs were prepared and directly used for amplified DNA detection without further modifications. To overcome the incompatibility of the synthesized Chit-AuNPs solutions with the LAMP reaction, we immobilized appropriate amounts of Chit-AuNPs inside the lid of the tubes by surface tension, which was mixed with the reaction after the amplification process was completed. Using the abovementioned one-tube assay, we avoided possible aerosol contamination by eliminating the need to open the tube and developed an endpoint colorimetric assay with a time-to-result of ∼35 min. Based on the presence or absence of free Chit in the Chit-AuNPs suspension, we demonstrated two colorimetric detection approaches, both of which could be used in the presence of crude saliva samples. Moreover, we showed that both methods can be used for the efficient naked-eye detection of the *Salmonella* target in the range of 5-1000 cfu/reaction, while the assay with the optimized free Chit exhibited an impressive detection limit of 1 cfu/reaction in both pure and crude saliva samples. In addition to the amplification and detection of *Salmonella*, the SARS-CoV-2 target was used as a proof of the method’s generic applicability toward detecting viral targets in combination with reverse transcriptase LAMP (RT-LAMP), giving a detection limit of 10 copies/reaction. Effective detection of both the aforementioned targets (bacterial-*Salmonella* and viral-SARS-CoV-2) is critical in reducing and monitoring possible outbreaks and providing fast diagnostic tools for application in healthcare and agri-food safety.

## Results and Discussion

### Synthesis and characterization of Chit-AuNPs

The Chit-AuNPs “green” synthesis employed in this study follows a chemical reduction process, where chitosan has a double role of a reducing and capping/stabilizing agent. Upon protonation of their amine groups in the presence of acetic acid, chitosan chains electrostatically adsorb negatively charged Au (III) ions that are reduced to neutral Au atoms, and further agglomerate to form AuNPs capped with chitosan.^37^ A schematic representation of the reaction is shown in Fig. 1a. We used three different Chit concentrations (0.15%, 0.25% and 0.35%, w/v) to prepare the final colloidal solutions, which presented different shades of red color, indicating the efficient Chit-AuNPs synthesis in different sizes and concentrations.

**Figure 1:**
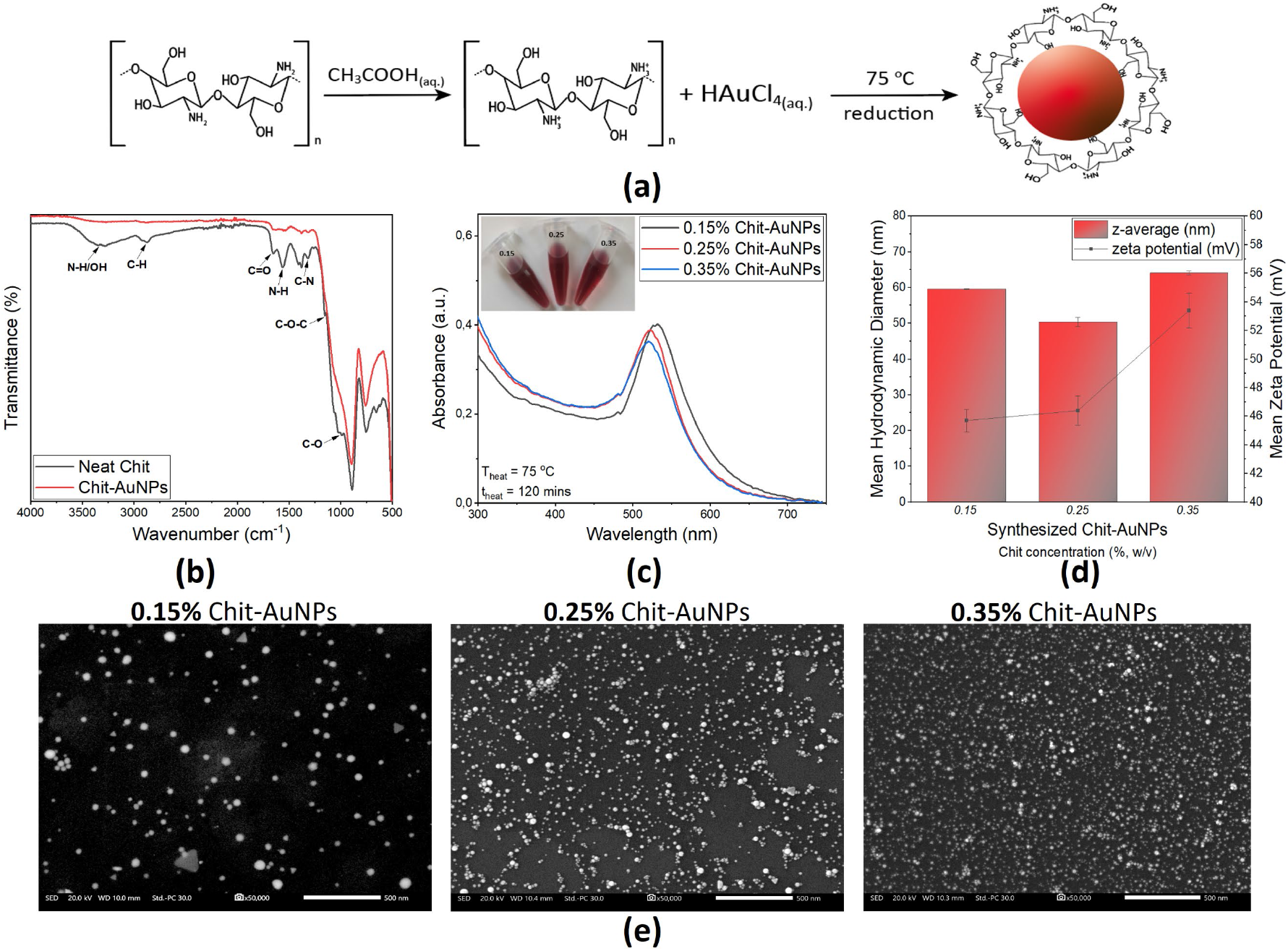
a) Schematic representation of AuNPs formulation using Chit dissolved in 1% (v/v) aqueous acetic acid. b) Representative ATR-FTIR spectra of neat Chit (black line) and Chit-AuNPs (red line), along with the peaks of interest. c) UV-Vis absorbance spectra, d) DLS and ZP measurements and e) SEM images (500 nm scale bar), of the Chit-AuNPs synthesized with different Chit concentrations (0.15%, 0.25% and 0.35%, w/v) at 75°C for 2h.

Attenuated total reflectance-Fourier transform infrared (ATR-FTIR) spectroscopy was conducted for neat Chit and Chit-AuNPs samples to further study the interaction between the Chit polymer and the formed AuNPs. The characteristic absorption bands from the functional groups of neat Chit observed in Fig. 1b were identified based on the literature.^38^ When comparing the neat Chit graph with the Chit-AuNPs one, a decrease in the intensity of several peaks, as well as shifts to lower energies were observed in the latter case. Based on these results, a contact between Chit and AuNPs through different chemical groups can be concluded (supramolecular interaction), in agreement with previous reports.^39,40^

As shown in the absorption spectra of the synthesized Chit-AuNPs (Fig. 1c), all the solutions have SPR bands, with the SPR peaks (λ_max_) at 532, 523 and 520 nm for 0.15%, 0.25% and 0.35% (w/v) Chit-AuNPs, respectively. This implies an inversely proportional relation between the Chit concentration and the synthesized Chit-AuNPs sizes, in agreement with similar studies.^41,42^ Regarding the full width at half maximum (FWHM), it is larger in the lowest Chit concentration, implying bigger polydispersity in the Chit-AuNPs sizes.^43^ SEM images further confirm the above (Fig. 1e), wherein the Chit-AuNPs synthesized with the lowest Chit concentration comprise larger and more polydisperse NPs. Besides spherical ones, thin triangular prismatic AuNPs were observed, particularly in the Chit-AuNPs synthesized with the lowest Chit concentration, possibly due to the slower AuNPs growth process.^44^ Based on the SEM analysis, the calculated mean Chit-AuNPs sizes were 38.4, 13.8 and 10.7 nm, while their calculated concentrations were 0.57, 12.3 and 26.4 nM for the 0.15%, 0.25% and 0.35% (w/v) Chit concentration used for the AuNPs synthesis, respectively (detailed calculation described in the ESI).

Dynamic light scattering (DLS) and zeta potential (ZP) measurements revealed a mean hydrodynamic diameter (D_h_) of 59.6, 50.4 and 64.1 nm, polydispersity index values (PDI) at around 0.46, 0.42 and 0.29 and mean z-potential values of +45.7, +46.4 and +53.4 mV for the Chit-AuNPs synthesized using 0.15%, 0.25% and 0.35% (w/v) Chit, respectively (Fig. 1d). In general, the calculated D_h_ is higher than the NPs size calculated via microscopy techniques, owing to the hydration shell around the NPs affecting their diffusion speed, and by extension their calculated size.^45^ Regarding the PDI, it is higher in the Chit-AuNPs solution synthesized with the lowest Chit concentration (0.15%, w/v), in agreement with the abovementioned conclusions based on the FWHM of the UV-Vis spectra. Z-potential values are positive owing to the cationic nature of Chit in the acidic solution, with values greater than 25 mV, implying a high degree of stability for all the colloidal solutions. Notably, the above values are also affected by the presence of free dissolved Chit in the solutions, that may undergo self-crosslinking to produce Chit polymer chains.^45^

### Naked-eye detection of LAMP amplicons in purified samples using Chit-AuNPs

A previous study has shown that directly incorporating water-soluble Chit in a LAMP reaction inhibits DNA amplification because positively charged Chit conjugates with negatively charged DNA or interferes with LAMP primer annealing.^36^ This was further confirmed when we added either dissolved neat Chit, or Chit-AuNPs in their originally synthesized acidic environment (replaced 5μL of nuclease-free water with Chit or Chit-AuNPs) and included as well 0.5 μL of LAMP fluorescent dye (readable in the SYBR^®^/FAM channel) for detection; in both cases, no amplification signal was observed (Fig. S1), as the pH of the final mixture (measured with a pH strip) is ∼4.5 owing to the acetic acid addition.

Based on the above, we developed a single-tube assay where 7.5 μL of the synthesized colloidal Chit-AuNPs solutions were immobilized, including as well free dissolved non-reacted Chit, inside the lids of the tubes (Fig. 2a). This approach was used to test all three different-sized Chit-AuNPs, employing *Salmonella* InvA gene as the target, within a concentration range of 1 - 1000 cfu/reaction. After the LAMP reactions were completed (30 min), a brief spin-down (∼10 s) of the tubes was carried out in order to mix the solutions.

**Figure 2:**
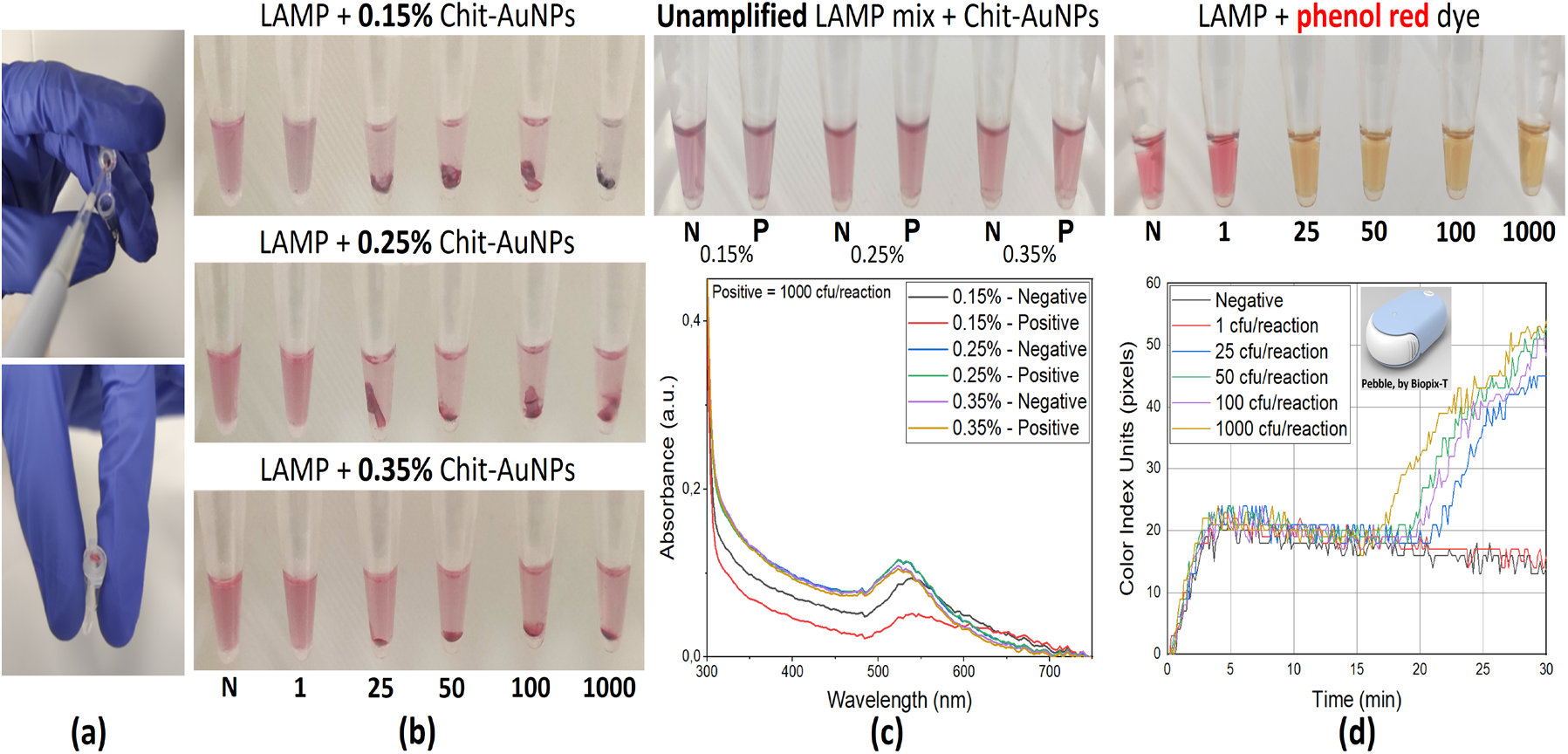
a) Immobilizing appropriate amounts of Chit-AuNPs solutions inside the lids of the tubes. b) Endpoint colorimetric results in purified samples, using the Chit-AuNPs synthesized with the different Chit concentrations; pellet formation and supernatant discoloration can be observed with increasing the number of target cfu/reaction. c) Photograph of the tubes and UV-Vis absorbance spectra of unamplified LAMP reaction mixed with the different Chit-AuNPs solutions. d) Photograph of the tubes & real-time qcLAMP diagram, using phenol red (pH indicator). Note: The values of 1-1000 below the photographs of the 0.2 mL tubes correspond to Salmonella target concentration in cfu/reaction, N corresponds to negative samples, and P to positives with 1000 cfu/reaction. Experiments were performed in triplicate.

According to Fig. 2b, in all cases (0.15%, 0.25% and 0.35% (w/v) Chit-AuNPs), the 25 cfu/reaction *Salmonella* concentration was detected based on pellet creation and supernatant discoloration, while the negative samples were stable. Moreover, in the Chit-AuNPs synthesized with the lowest Chit concentration (0.15%, w/v), a slight color change was observed even at 1 cfu/reaction *Salmonella* concentration. The higher sensitivity observed for the 0.15% (w/v) Chit-AuNPs was further confirmed by directly adding the three different Chit-AuNPs solutions inside a LAMP reaction mixture with 1000 cfu/reaction (positive) or without (negative) target DNA, and observing the color change without amplification. As shown in Fig. 2c, for the Chit-AuNPs synthesized with the lowest Chit concentration (0.15%, w/v), a color change was detected by the naked-eye between the negative and the positive sample, along with a drop in the UV-Vis absorbance intensity in the case of the positive sample. In the case of the Chit-AuNPs synthesized with the higher Chit concentrations, both the negative and positive samples retained the same color and absorbance intensities.

Parallel to the abovementioned endpoint naked-eye colorimetric detection using Chit-AuNPs, colorimetric LAMP using phenol red (pH indicator) was used to further evaluate the assay. For these experiments, we used a real-time colorimetric device, which can perform quantitative colorimetric LAMP (qcLAMP).^46^ As shown in Fig. 2d, we could also detect down to 25 cfu/reaction *Salmonella* concentration using this method, but not the 1 cfu/reaction. The time-to-positive result was ∼17 min for the 1000 cfu/reaction, ∼19.5 min for the 100 and 50 cfu/reaction and ∼21 min for the 25 cfu/reaction. These results confirm that the Chit-AuNPs colorimetric detection methodology is equally or more sensitive to a colorimetric real time method, although the Chit-AuNPs approach cannot be used for real-time quantification.

### Naked-eye detection of LAMP amplicons in saliva samples using Chit-AuNPs

Experiments were also conducted with crude saliva samples to further test the applicability of the Chit-AuNPs-based colorimetric assay for POC applications. Initially, we tested the effect of saliva on the stability of the Chit-AuNPs in the LAMP reaction by testing one positive, containing 100 cfu/reaction of the *Salmonella* target, and one negative sample. In particular, we replaced 2.5 or 5 μL of the nuclease-free water in the LAMP mix with lysed saliva. These amounts correspond to a final 10% and 20% saliva sample concentration, respectively, one of the highest % of crude samples in a LAMP reported so far. At the same time, similar amounts (7.5 μL) of the three Chit-AuNPs solutions were immobilized inside the lids of the tubes. After the LAMP reactions were completed, a brief spin-down of the tubes was conducted to mix the solutions and help reveal the changes in the color fast. A pellet was observed in the negative samples, primarily at the two lower concentrations (0.15% and 0.25%, w/v), along with an intense discoloration of these two solutions (Fig. 3a, left). After a second spin-down, the only solution that retained its color was the negative sample with 0.35% (w/v) Chit-AuNPs (Fig. 3a, right) for both 2.5 and 5 μL saliva samples, while the discoloration of the negatives (0.15% and 0.25%, w/v) was stronger and faster in the 5 μL (20%) than that of the 2.5 μL (10%) saliva sample. The positive samples containing 100 cfu/reaction of the target presented aggregation in all cases, as observed before with pure samples (Fig. 2b). Out of these results, it can be concluded that in the negative samples, the free Chit was not sufficient to efficiently protect the 0.15% and 0.25% (w/v) Chit-AuNPs from aggregation induced by the different saliva components. However, we cannot exclude the effect of the three different Chit-AuNPs in the above behavior because of the different Chit-AuNPs sizes and concentrations.

**Figure 3:**
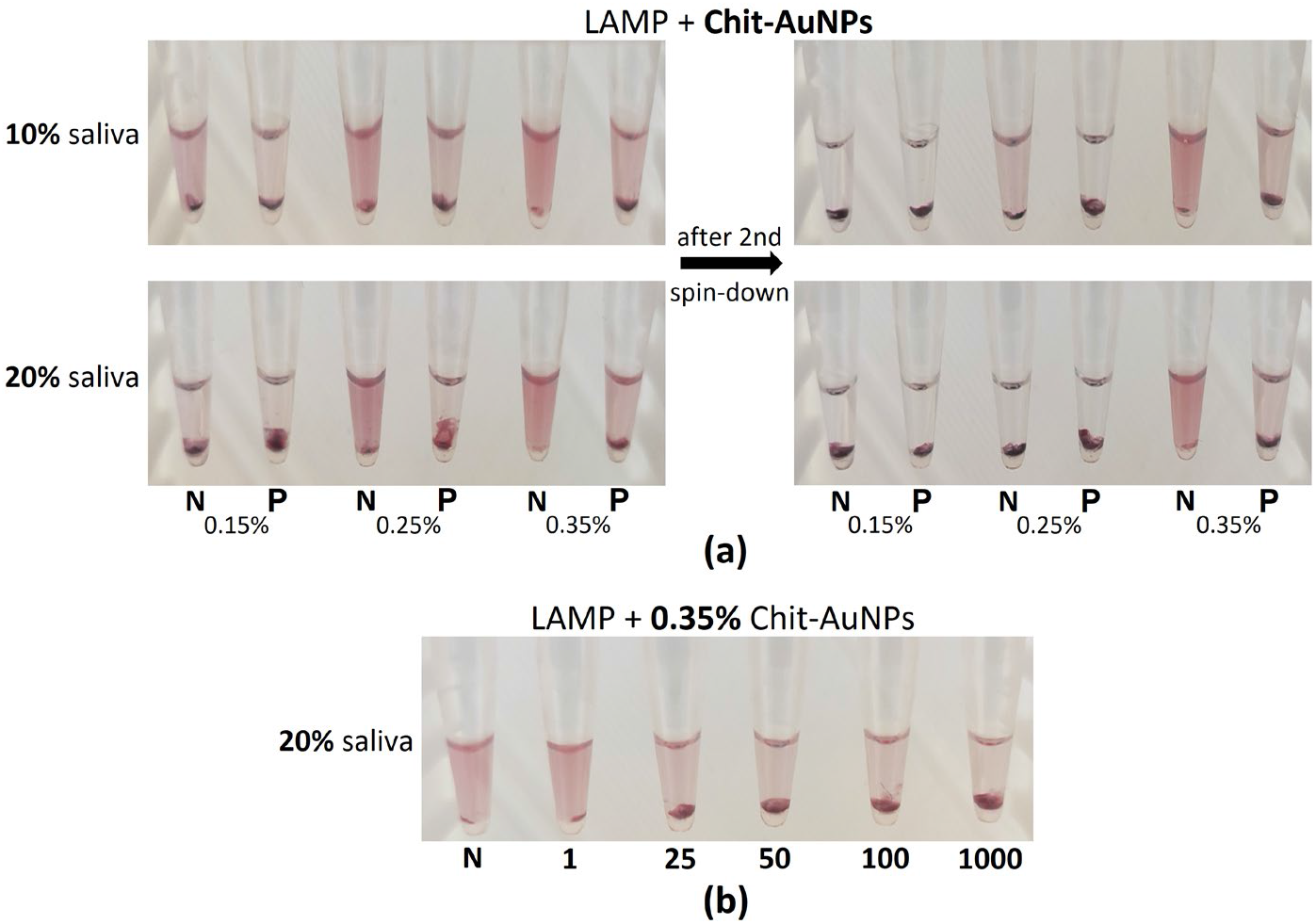
a) Observation of the different Chit-AuNPs stabilization efficiency on the negative samples in the presence of different amounts of lysed saliva samples inside the LAMP mix. b) Endpoint colorimetric results using the synthesized 0.35% (w/v) Chit-AuNPs, in different target cfu/reaction, in a LAMP mix containing 5 μL saliva (20%). Note: The values of 1-1000 below the 0.2 mL tubes correspond to Salmonella target concentration in cfu/reaction, N corresponds to negative samples, and P to positives with 100 cfu/reaction. Experiments were performed in triplicate.

The solution of 0.35% (w/v) Chit-AuNPs, the most efficient to maintain Chit-AuNPs stabilization in the presence of saliva, was further used for the clear naked-eye detection of different target amounts within the range of 25-1000 cfu/reaction of the *Salmonella* target (Fig. 3b).

To further evaluate the effect of free Chit and the final solution’s pH on differentiating negative and positive samples, 0.5 mL of 0.15% (w/v) Chit-AuNPs solution was centrifuged, as it had the worst performance in the saliva samples. Afterward, the supernatant containing the free Chit was removed, and the Chit-AuNPs pellets were redispersed in three solutions: (a) 1% (v/v) aqueous acetic acid; (b) ultrapure water; and (c) 1% (v/v) aqueous acetic acid containing 0.15% (w/v) Chit. The resulting solutions showed no aggregation after the purification step and redispersion in the different media (Fig. S2).

As seen before, the initially synthesized 0.15% (w/v) Chit-AuNPs were not able to provide a distinct colorimetric difference between the positive (100 cfu/reaction) and negative samples after the LAMP reaction in 20% saliva, because Chit-AuNPs aggregation occurred in both solutions (Fig. 4a, middle). The addition of excess Chit in the solution (0.15%, w/v) was able to inhibit the complete Chit-AuNPs aggregation of negative samples (Fig. 4a, right), similar to 0.35% (w/v) Chit-AuNPs solution in the crude saliva samples, further confirming the role of the free Chit itself in the solution. Interestingly, the Chit-AuNPs solution without any free Chit, i.e., the one redispersed in 1% (v/v) aqueous acetic acid, provided the opposite results, with aggregation in the negative sample and the positive ones remained stable (Fig. 4a, left). When we repeated these experiments in pure samples, a distinct colorimetric difference was observed in all cases (Fig. 4b).

**Figure 4:**
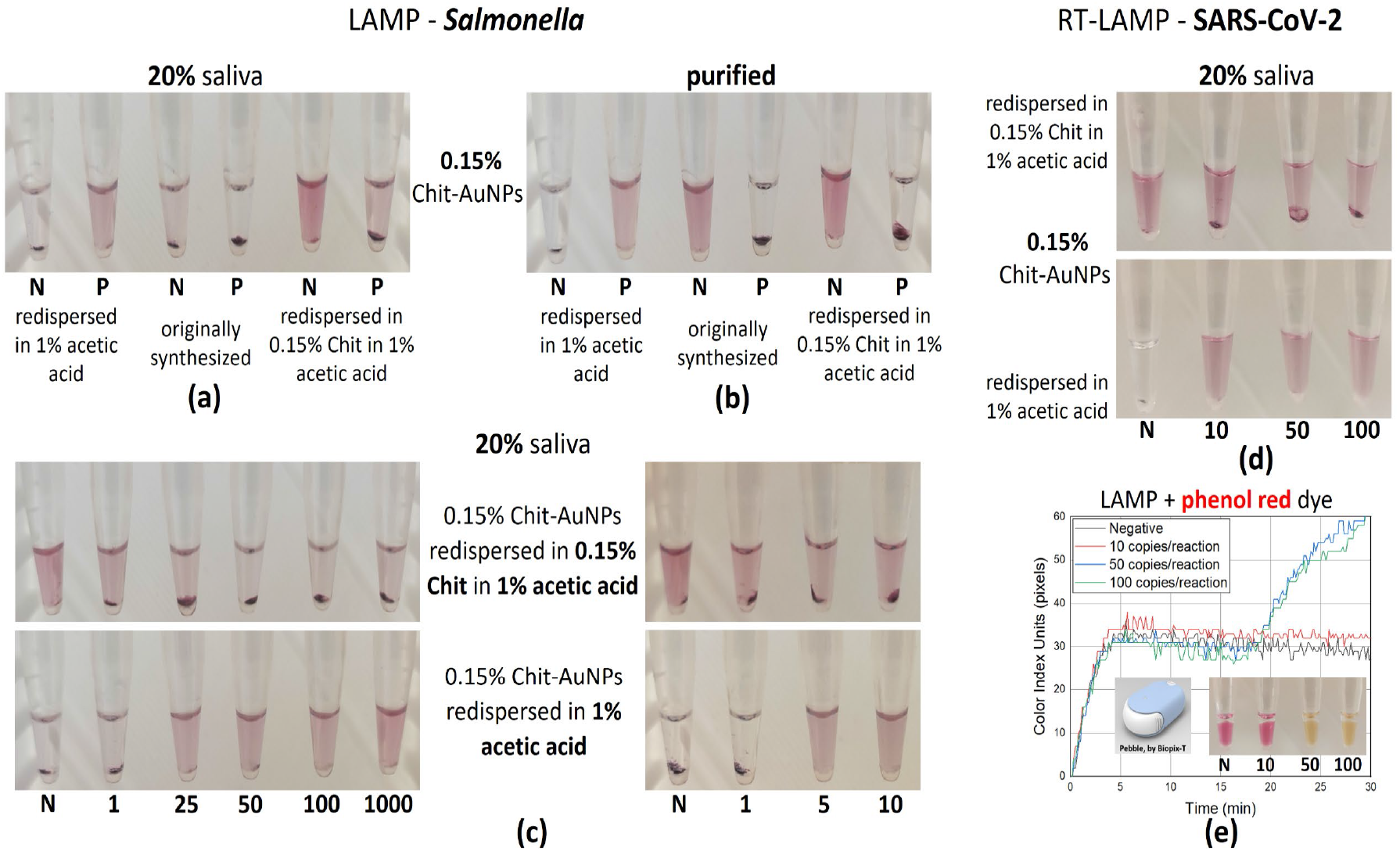
a, b) Comparative colorimetric results between the initially synthesized 0.15% (w/v) Chit-AuNPs, and the redispersed Chit-AuNPs in either 1% (v/v) aqueous acetic acid or 0.15% (w/v) Chit in 1% (v/v) aqueous acetic acid for both purified and crude saliva samples (20%). c) Endpoint colorimetric results using the redispersed Chit-AuNPs – mirror-like results based on the presence or absence of free Chit in the solution, for 20% saliva samples (Salmonella target). d, e) Photographs of endpoint colorimetric detection and real-time qcLAMP diagram, after RT-LAMP reaction containing 20% saliva samples (SARS-CoV-2 target). Note: The values of 1-1000 below the 0.2 mL tubes correspond to target cfu (Salmonella) or copies (SARS-CoV-2)/reaction, N corresponds to negative samples, and P to positives with 100 cfu/reaction. Experiments were performed in at least triplicate.

Moreover, the two 0.15% (w/v) Chit-AuNPs solutions, with or without excess free Chit in 1% (v/v) aqueous acetic acid, were tested with different amounts of the target within the range of 1-1000 cfu/reaction (Fig. 4c, left) for the saliva samples. In both cases, the 25 cfu/reaction target concentration was easily detected colorimetrically by naked-eye in the LAMP reaction, while the 1 cfu/reaction target concentration was detected only when the Chit-AuNPs were redispersed in excess Chit (0.15%, w/v). Testing closer the efficiency of the two assays in the range of 1 and 25 cfu/reaction confirmed the abovementioned observations; notably, the 1 cfu/reaction target concentration was detected in half of the tested samples (number of samples: 6) (Fig. 4c, right), probably reflecting the probability of capturing the target with every pipetted sampling.

Finally, RT-LAMP experiments further confirmed that the aforementioned method could also detect SARS-CoV-2 in the presence of 20% saliva, with a detection limit of 10 copies/reaction (Fig. 4d). This result proves that the assay is flexible and generic as it can be used with different targets and in combination with RT. Comparison with colorimetric detection using phenol red (qcLAMP) further confirmed the higher sensitivity of the Chit-AuNPs detection method, compared to pH sensitive dyes. (Fig. 4e). The time-to-positive was ∼19 min for both 100 and 50 cfu/reaction target concentration in the phenol red system, while the 10 copies/reaction were not able to be detected, probably due to insufficient H^+^ by-products produced during amplification that shall induce a significant pH drop, and/or due to the possibly increased buffer capacity and pH in the presence of the crude (5 μL saliva) sample.

### Mechanism of the Chit-AuNPs-based colorimetric detection of LAMP amplicons

To explain the abovementioned results, we investigated both the ability of Chit to exist in a pH-dependent charged form and the mechanism behind AuNPs stabilization. For a pKa of ∼6.5, Chit is protonated at a lower pH and exists in a cationic form and a DNA-binding state. AuNPs can be stabilized and remain dispersed in a solution because of electrostatic repulsive forces. Additionally, when polymer molecules are attached to the AuNPs, forming a coating, steric stabilization is achieved via repulsive forces that separate the particles from one another, while depletion stabilization can also be observed in the presence of free polymeric molecules in the solution. The combination of all three colloidal stabilization factors is shown to provide ultrahigh NPs stability under different conditions,^47,48^ which would otherwise lead to aggregation. Herein, different destabilization agents (salt, non-target DNA, dNTPs, and other elements in the saliva) in the LAMP mix can lead to NPs aggregation.

Based on the above, our results indicate that when using Chit-AuNPs with free Chit in purified negative samples, the free Chit polymer can protect the Chit-AuNPs from aggregation induced by the different LAMP reagents, keeping the AuNPs dispersed in the solution owing to the depletion stabilization effects in addition to the Chit-AuNPs electrosteric (electrostatic and steric) stabilization mechanism (Fig. 2b). However, it appears that there is an optimum free Chit amount required for stabilization depending on the sample-type (purified vs saliva). Although the free Chit in the 0.15% (w/v) synthesized Chit-AuNPs is sufficient to provide stabilization in purified negative samples (Fig. 4b, middle), it is not equally effective in crude negative samples (Fig. 4a, middle). In saliva, this amount was insufficient to cope with the extra DNA released upon cell-lysis and possible variations in the pH and salt;^49,50^ hence, a higher amount of free Chit concentration is essential to induce the same result. Alternatively, free Chit operates as a “buffer” solution providing enhanced stability of the Chit-AuNPs in both pure and saliva-containing negative samples. In both pure and saliva-containing positive samples, the high yields of negatively charged DNA amplicons combined with the positively charged free Chit and Chit-AuNPs (in the acidic final solution of ∼4.5 pH) disturb stabilization via electrostatic interactions, causing Chit-AuNPs aggregation. However, in the presence of saliva, the negative samples also tend to aggregate after ∼4 days, possibly due to the differently charged antagonistic populations inside the solution (Fig. S3a). A schematic diagram of the Chit-AuNPs stabilization mechanism with free Chit is depicted in Fig. 5, top line.

**Figure 5:**
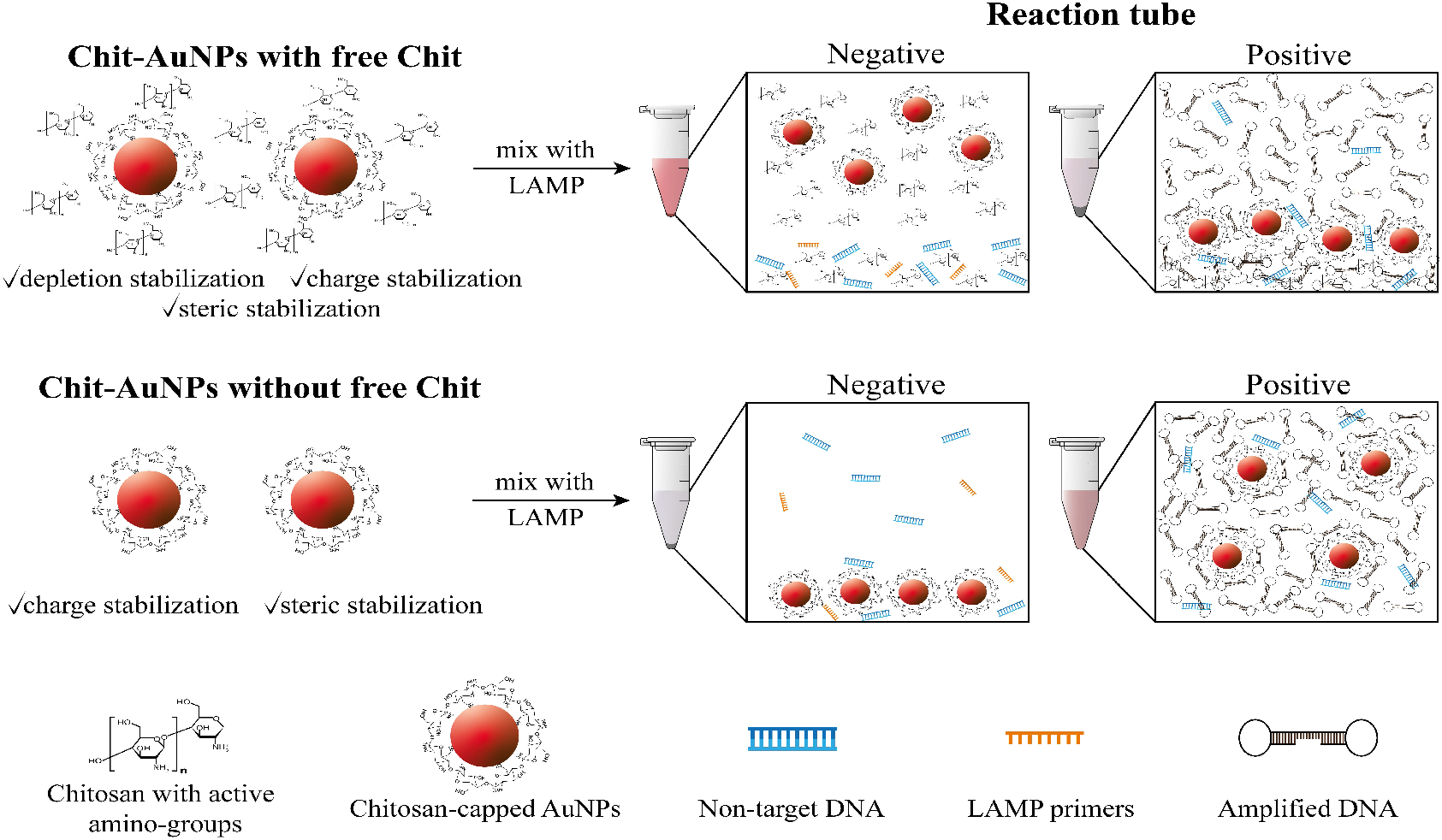
Close-up image of the Chit-AuNPs, free Chit and (amplified) DNA interactions inside the LAMP mix after amplification reaction. Note that the Chit-AuNPs solutions are dissolved in 1% (v/v) aqueous acetic acid in both cases.

Conversely, when using Chit-AuNPs without free Chit in the solution, a “mirror effect” is observed; the negative samples are destabilized in the presence of the LAMP mix even in the presence of 20% saliva, with the Chit-AuNPs aggregating due to the absence of the free Chit depletion protection. At the same time, the positive samples remain stable, as the large amounts of DNA amplicons can electrostatically coat the positively charged Chit-AuNPs, protecting them from aggregation via repulsion forces (Fig. 5, bottom line). The complete coating/stabilization of the positively charged Chit-AuNPs with negatively charged amplified DNA in the stable positive samples was also confirmed by the z-potential measurements (Fig. S4a), where the solution showed a negative z-value (∼ -7.1 mV). This was contrary to the stable negative samples of the free Chit method, where the solution presented a positive z-value (∼ +10.7 mV), owing to the dominating presence of free Chit stabilizing the solution. UV-Vis measurements further proved the efficient conjugation of Chit-AuNPs with DNA in the neat Chit-AuNPs method (no free Chit) via the observed red-shift of SPR λ_max_ from 532 to 541 nm and a drop in the absorbance intensity (Fig. S4b). The low pH of ∼4.5 in the final solution (LAMP mix + Chit-AuNPs redispersed in 1% (v/v) aqueous acetic acid) is essential for the electrostatic stabilization of the positive samples; indeed, when repeating the same experiment with Chit-AuNPs redispersed in ultrapure water, the positive samples also aggregated (Fig. S3b). In ultrapure water, the pH of the final solution is close to the pKa of Chit (∼6.5), which partially neutralizes the charge of the Chit polymer, reducing the electrostatic stabilization or repulsive Coulomb forces between the Chit-AuNPs, leaving only the steric stabilization provided by the Chit coating of the AuNPs. Consequently, the positive samples also tend to aggregate in the LAMP mix owing to the absence of strong electrostatic interactions with the negatively charged DNA. Notably, the concentration of Chit-AuNPs used in this method is also crucial, as low concentrations of NPs may lead to stabilization by the non-target DNA contained in the saliva, eliminating the possibility of differentiating the negative samples from the positive ones (false positives).

## Conclusions

Herein, we exploited the effect of free Chit in synthesized Chit-AuNPs solutions to create two different naked-eye endpoint colorimetric assays combined with LAMP. Using the different stabilization forces (depletion, electrostatic, and steric) between the pH-responsive Chit and Chit-AuNPs, we developed two mirror-like assays, one in the presence and the other in the absence of free Chit. In both cases, we demonstrated the ultrasensitive eye-detection of the *Salmonella* target with a detection limit of 1 cfu/reaction (40 cfu/mL) in the presence of free Chit and 5 cfu/reaction (200 cfu/mL) in the absence of free Chit in the Chit-AuNPs solution. The above performance was also shown in an impressive final saliva concentration of 20%. The general applicability of the method toward the detection of viral targets was also demonstrated when 10 copies/reaction (400 copies/mL) of SARS-CoV-2 in 20% saliva, amplified with RT-LAMP were successfully detected by naked-eye. This colorimetric detection method combines many attractive features, such as simplicity, high sensitivity, and rapid results, making it an ideal candidate for incorporation in POC applications. Using a “green” synthesis of Chit-AuNPs is considered an added advantage. Being a target-independent method, this assay does not provide any added degree of specificity, which entirely depends on the selected primers. However, this limitation leads to one advantage of this method, which is the compatibility with any kind of (RT-)LAMP assay. Currently, both assays rely on endpoint detection; however, the assay using free Chit and Chit-AuNPs could be employed for quantitative results based on the decrease of the absorbance intensity of the supernatant (discoloration), when the final solutions are measured at the same time. Such a quantification method could be more sensitive than colorimetric assays using pH-sensitive dyes, because the free Chit/Chit-AuNPs system essentially detects DNA under appropriate pH conditions and not LAMP-induced pH changes. Finally, this assay could possibly be applied to other isothermal amplification methods, such as (RT-)RPA, where amplification happens at even lower temperatures (37-42 °C), providing another efficient, fast, sensitive and cost-effective POC system.

## Data Availability

All data produced in the present study are available upon reasonable request to the authors.

## Associated Content

### Supporting Information

Reagents and materials, experimental details about Chit-AuNPs synthesis, purification and characterization, theoretical calculation for the Chit-AuNPs molar concentration, experimental details for the colorimetric LAMP assays preparation and evaluation.

Supplementary Figures; S1: LAMP reaction in the presence of Chit-AuNPs, S2: Purification of Chit-AuNPs via centrifugation, S3: Effect of time and pH on the stability of the final solutions, S4: Zeta-Potential and UV-Vis Absorbance measurements of the final stable solutions.

## Author Information

### Author Contributions

**S.G**. conceived and performed the experimental work, as well as wrote the first draft of the manuscript. **I.S**. carried out experimental work and discussed the results. **E.G**. supervised the whole work, discussed experiments and results and wrote the final manuscript. All authors have given approval to the final version of the manuscript.

## Acknowledgments

This work has received funding from the EC through the HORIZON2020 FET-OPEN-2018-2020 grant No 862840 (project acronym “FREE@POC”).

The authors would like to acknowledge Prof. M. Stylianakis and Dr. G. Kenanakis (IESL-FORTH) for providing access to the DLS and FTIR instruments, respectively. Ms. K. Katsara and Ms. A. Manousaki (IESL-FORTH) are also acknowledged for their assistance in taking FTIR measurements and SEM images, respectively.

## References

(1) Heithoff, D. M.; Barnes, L.; Mahan, S. P.; Fox, G. N.; Arn, K. E.; Ettinger, S. J.; Bishop, A. M.; Fitzgibbons, L. N.; Fried, J. C.; Low, D. A.; Samuel, C. E.; Mahan, M. J. Assessment of a Smartphone-Based Loop-Mediated Isothermal Amplification Assay for Detection of SARS-CoV-2 and Influenza Viruses. JAMA Netw Open 2022, 5 (1), e2145669. https://doi.org/10.1001/jamanetworkopen.2021.45669.

(2) Garg, N.; Sahu, U.; Kar, S.; Ahmad, F. J. Development of a Loop-Mediated Isothermal Amplification (LAMP) Technique for Specific and Early Detection of Mycobacterium Leprae in Clinical Samples. Sci Rep 2021, 11 (1), 9859. https://doi.org/10.1038/s41598-021-89304-2.

(3) Narushima, J.; Kimata, S.; Soga, K.; Sugano, Y.; Kishine, M.; Takabatake, R.; Mano, J.; Kitta, K.; Kanamaru, S.; Shirakawa, N.; Kondo, K.; Nakamura, K. Rapid DNA Template Preparation Directly from a Rice Sample without Purification for Loop-Mediated Isothermal Amplification (LAMP) of Rice Genes. Biosci Biotechnol Biochem 2020, 84 (4), 670–677. https://doi.org/10.1080/09168451.2019.1701406.

(4) Kaur, A.; Kapil, A.; Elangovan, R.; Jha, S.; Kalyanasundaram, D. Highly-Sensitive Detection of Salmonella Typhi in Clinical Blood Samples by Magnetic Nanoparticle-Based Enrichment and in-Situ Measurement of Isothermal Amplification of Nucleic Acids. PLoS One 2018, 13 (3), e0194817. https://doi.org/10.1371/journal.pone.0194817.

(5) Österdahl, M. F.; Lee, K. A.; Lochlainn, M. N.; Wilson, S.; Douthwaite, S.; Horsfall, R.; Sheedy, A.; Goldenberg, S. D.; Stanley, C. J.; Spector, T. D.; Steves, C. J. Detecting SARS-CoV-2 at Point of Care: Preliminary Data Comparing Loop-Mediated Isothermal Amplification (LAMP) to Polymerase Chain Reaction (PCR). BMC Infect Dis 2020, 20 (1), 783. https://doi.org/10.1186/s12879-020-05484-8.

(6) Das, D.; Lin, C.-W.; Chuang, H.-S. LAMP-Based Point-of-Care Biosensors for Rapid Pathogen Detection. Biosensors (Basel) 2022, 12 (12), 1068. https://doi.org/10.3390/bios12121068.

(7) Garrido-Maestu, A.; Prado, M. Naked-eye Detection Strategies Coupled with Isothermal Nucleic Acid Amplification Techniques for the Detection of Human Pathogens. Compr Rev Food Sci Food Saf 2022, 21 (2), 1913–1939. https://doi.org/10.1111/1541-4337.12902.

(8) Charoenpanich, P.; Mungkung, A.; Seeviset, N. A PH Sensitive, Loop-Mediated Isothermal Amplification Assay for Detection of Salmonella in Food. Research Article Science, Engineering and Health Studies 2020 (3), 160–168.

(9) Brown, T. A.; Schaefer, K. S.; Tsang, A.; Yi, H. A.; Grimm, J. B.; Lemire, A. L.; Jradi, F. M.; Kim, C.; McGowan, K.; Ritola, K.; Armstrong, D. T.; Mostafa, H. H.; Korff, W.; Vale, R. D.; Lavis, L. D. Direct Detection of SARS-CoV-2 RNA Using High-Contrast PH-Sensitive Dyes. J Biomol Tech 2021, 32 (3), 121–133. https://doi.org/10.7171/jbt.21-3203-007.

(10) Zasada, A. A.; Wiatrzyk, A.; Czajka, U.; Brodzik, K.; Formińska, K.; Mosiej, E.; Prygiel, M.; Krysztopa-Grzybowska, K.; Wdowiak, K. Application of Loop-Mediated Isothermal Amplification Combined with Colorimetric and Lateral Flow Dipstick Visualization as the Potential Point-of-Care Testing for Corynebacterium Diphtheriae. BMC Infect Dis 2020, 20 (1), 308. https://doi.org/10.1186/s12879-020-05037-z.

(11) Tanner, N. A.; Zhang, Y.; Evans, T. C. Visual Detection of Isothermal Nucleic Acid Amplification Using PH-Sensitive Dyes. Biotechniques 2015, 58 (2), 59–68. https://doi.org/10.2144/000114253.

(12) Daniel, M.-C.; Astruc, D. Gold Nanoparticles: Assembly, Supramolecular Chemistry, Quantum-Size-Related Properties, and Applications toward Biology, Catalysis, and Nanotechnology. 2004. https://doi.org/10.1021/cr030698.

(13) Jazayeri, M. H.; Aghaie, T.; Avan, A.; Vatankhah, A.; Ghaffari, M. R. S. Colorimetric Detection Based on Gold Nano Particles (GNPs): An Easy, Fast, Inexpensive, Low-Cost and Short Time Method in Detection of Analytes (Protein, DNA, and Ion). Sens Biosensing Res 2018, 20, 1–8. https://doi.org/10.1016/j.sbsr.2018.05.002.

(14) Teixeira, A.; Paris, J. L.; Roumani, F.; Diéguez, L.; Prado, M.; Espiña, B.; Abalde-Cela, S.; Garrido-Maestu, A.; Rodriguez-Lorenzo, L. Multifuntional Gold Nanoparticles for the SERS Detection of Pathogens Combined with a LAMP–in–Microdroplets Approach. Materials 2020, 13 (8), 1934. https://doi.org/10.3390/ma13081934.

(15) Arunrut, N.; Kampeera, J.; Suebsing, R.; Kiatpathomchai, W. Rapid and Sensitive Detection of Shrimp Infectious Myonecrosis Virus Using a Reverse Transcription Loop-Mediated Isothermal Amplification and Visual Colorogenic Nanogold Hybridization Probe Assay. J Virol Methods 2013, 193 (2), 542–547. https://doi.org/10.1016/j.jviromet.2013.07.017.

(16) Seetang-Nun, Y.; Jaroenram, W.; Sriurairatana, S.; Suebsing, R.; Kiatpathomchai, W. Visual Detection of White Spot Syndrome Virus Using DNA-Functionalized Gold Nanoparticles as Probes Combined with Loop-Mediated Isothermal Amplification. Mol Cell Probes 2013, 27 (2), 71–79. https://doi.org/10.1016/j.mcp.2012.11.005.

(17) Zhang, Y.; Chen, M.; Liu, C.; Chen, J.; Luo, X.; Xue, Y.; Liang, Q.; Zhou, L.; Tao, Y.; Li, M.; Wang, D.; Zhou, J.; Wang, J. Sensitive and Rapid On-Site Detection of SARS-CoV-2 Using a Gold Nanoparticle-Based High-Throughput Platform Coupled with CRISPR/Cas12-Assisted RT-LAMP. Sens Actuators B Chem 2021, 345, 130411. https://doi.org/10.1016/j.snb.2021.130411.

(18) Cao, Y.; Wu, J.; Pang, B.; Zhang, H.; Le, X. C. CRISPR/Cas12a-Mediated Gold Nanoparticle Aggregation for Colorimetric Detection of SARS-CoV-2. Chemical Communications 2021, 57 (56), 6871–6874. https://doi.org/10.1039/D1CC02546E.

(19) López-Valls, M.; Escalona-Noguero, C.; Rodríguez-Díaz, C.; Pardo, D.; Castellanos, M.; Milán- Rois, P.; Martínez-Garay, C.; Coloma, R.; Abreu, M.; Cantón, R.; Galán, J. C.; Miranda, R.; Somoza, Á.; Sot, B. CASCADE: Naked Eye-Detection of SARS-CoV-2 Using Cas13a and Gold Nanoparticles. Anal Chim Acta 2022, 1205, 339749. https://doi.org/10.1016/j.aca.2022.339749.

(20) Wong, J. K. F.; Yip, S. P.; Lee, T. M. H. Ultrasensitive and Closed-Tube Colorimetric Loop-Mediated Isothermal Amplification Assay Using Carboxyl-Modified Gold Nanoparticles. Small 2014, 10 (8), 1495–1499. https://doi.org/10.1002/smll.201302348.

(21) Qin, A.; Fu, L. T.; Wong, J. K. F.; Chau, L. Y.; Yip, S. P.; Lee, T. M. H. Precipitation of PEG/Carboxyl-Modified Gold Nanoparticles with Magnesium Pyrophosphate: A New Platform for Real-Time Monitoring of Loop-Mediated Isothermal Amplification. ACS Appl Mater Interfaces 2017, 9 (12), 10472–10480. https://doi.org/10.1021/acsami.7b00046.

(22) Ruang-areerate, T.; Saengsawang, N.; Ruang-areerate, P.; Ratnarathorn, N.; Thita, T.; Leelayoova, S.; Siripattanapipong, S.; Choowongkomon, K.; Dungchai, W. Distance-Based Paper Device Using Combined SYBR Safe and Gold Nanoparticle Probe LAMP Assay to Detect Leishmania among Patients with HIV. Sci Rep 2022, 12 (1), 14558. https://doi.org/10.1038/s41598-022-18765-w.

(23) Kong, C.; Wang, Y.; Fodjo, E. K.; Yang, G.; Han, F.; Shen, X. Loop-Mediated Isothermal Amplification for Visual Detection of Vibrio Parahaemolyticus Using Gold Nanoparticles. Microchimica Acta 2018, 185 (1), 35. https://doi.org/10.1007/s00604-017-2594-4.

(24) Mori, Y.; Hirano, T.; Notomi, T. Sequence Specific Visual Detection of LAMP Reactions by Addition of Cationic Polymers. BMC Biotechnol 2006, 6 (1), 3. https://doi.org/10.1186/1472-6750-6-3.

(25) Tammam, S. N.; Khalil, M. A. F.; Abdul Gawad, E.; Althani, A.; Zaghloul, H.; Azzazy, H. M. E. Chitosan Gold Nanoparticles for Detection of Amplified Nucleic Acids Isolated from Sputum. Carbohydr Polym 2017, 164, 57–63. https://doi.org/10.1016/j.carbpol.2017.01.051.

(26) de Oliveira, A. C.; Sabino, R. M.; Souza, P. R.; Muniz, E. C.; Popat, K. C.; Kipper, M. J.; Zola, R. S.; Martins, A. F. Chitosan/Gellan Gum Ratio Content into Blends Modulates the Scaffolding Capacity of Hydrogels on Bone Mesenchymal Stem Cells. Materials Science and Engineering: C 2020, 106, 110258. https://doi.org/10.1016/j.msec.2019.110258.

(27) Bravo-Anaya, L. M.; Soltero, J. F. A.; Rinaudo, M. DNA/Chitosan Electrostatic Complex. Int J Biol Macromol 2016, 88, 345–353. https://doi.org/10.1016/j.ijbiomac.2016.03.035.

(28) Bravo-Anaya, L. M.; Fernández-Solís, K. G.; Rosselgong, J.; Nano-Rodríguez, J. L. E.; Carvajal, F.; Rinaudo, M. Chitosan-DNA Polyelectrolyte Complex: Influence of Chitosan Characteristics and Mechanism of Complex Formation. Int J Biol Macromol 2019, 126, 1037–1049. https://doi.org/10.1016/j.ijbiomac.2019.01.008.

(29) Majdi, H.; Salehi, R.; Pourhassan-Moghaddam, M.; Mahmoodi, S.; Poursalehi, Z.; Vasilescu, S. Antibody Conjugated Green Synthesized Chitosan-Gold Nanoparticles for Optical Biosensing. Colloid Interface Sci Commun 2019, 33, 100207. https://doi.org/10.1016/j.colcom.2019.100207.

(30) Yazid, H.; Yassin, A. M.; Ruslan, A. Z.; Alias, S. H.; Adnan, R.; Md Jani, A. M. Synthesis of Chitosan-Gold Nanoparticles for Drug Delivery. Adv Mat Res 2014, 896, 280–283. https://doi.org/10.4028/www.scientific.net/AMR.896.280.

(31) Sun, I.-C.; Na, J. H.; Jeong, S. Y.; Kim, D.-E.; Kwon, I. C.; Choi, K.; Ahn, C.-H.; Kim, K. Biocompatible Glycol Chitosan-Coated Gold Nanoparticles for Tumor-Targeting CT Imaging. Pharm Res 2014, 31 (6), 1418–1425. https://doi.org/10.1007/s11095-013-1142-0.

(32) Tripathy, S.; Chalana, A. K.; Talukdar, A.; Rajesh, P. v.; Saha, A.; Pramanik, G.; Ghosh, S. Limited-Resource Preparable Chitosan Magnetic Particles for Extracting Amplification-Ready Nucleic Acid from Complex Biofluids. Analyst 2022, 147 (1), 165–177. https://doi.org/10.1039/D1AN01150B.

(33) Gómez Pérez, A.; González-Martínez, E.; Díaz Águila, C. R.; González-Martínez, D. A.; González Ruiz, G.; García Artalejo, A.; Yee-Madeira, H. Chitosan-Coated Magnetic Iron Oxide Nanoparticles for DNA and RhEGF Separation. Colloids Surf A Physicochem Eng Asp 2020, 591, 124500. https://doi.org/10.1016/j.colsurfa.2020.124500.

(34) Bakshi, P. S.; Selvakumar, D.; Kadirvelu, K.; Kumar, N. S. Chitosan as an Environment Friendly Biomaterial – a Review on Recent Modifications and Applications. Int J Biol Macromol 2020, 150, 1072–1083. https://doi.org/10.1016/j.ijbiomac.2019.10.113.

(35) da Silva, A. B.; Rufato, K. B.; de Oliveira, A. C.; Souza, P. R.; da Silva, E. P.; Muniz, E. C.; Vilsinski, B. H.; Martins, A. F. Composite Materials Based on Chitosan/Gold Nanoparticles: From Synthesis to Biomedical Applications. Int J Biol Macromol 2020, 161, 977–998. https://doi.org/10.1016/j.ijbiomac.2020.06.113.

(36) Diaz, L.; Li, Y.; Jenkins, D. M. Chemical Stabilization of Dispersed Escherichia Coli for Enhanced Recovery with a Handheld Electroflotation System and Detection by Loop-Mediated Isothermal AMPlification. PLoS One 2021, 16 (1), e0244956. https://doi.org/10.1371/journal.pone.0244956.

(37) Lakshmi Narayanan, R.; Sivakumar, M. Preparation and Characterization of Gold Nanoparticles in Chitosan Suspension by One-Pot Chemical Reduction Method. Nano Hybrids 2014, 6, 47–57. https://doi.org/10.4028/www.scientific.net/NH.6.47.

(38) Fernandes Queiroz, M.; Melo, K.; Sabry, D.; Sassaki, G.; Rocha, H. Does the Use of Chitosan Contribute to Oxalate Kidney Stone Formation? Mar Drugs 2014, 13 (1), 141–158. https://doi.org/10.3390/md13010141.

(39) Silva, A. T. B.; Coelho, A. G.; Lopes, L. C. da S.; Martins, M. V. A.; Crespilho, F. N.; Merkoçi, A.; Silva, W. C. da. Nano-Assembled Supramolecular Films from Chitosan-Stabilized Gold Nanoparticles and Cobalt(II) Phthalocyanine. J Braz Chem Soc 2013, 24 (8), 1237–1245. https://doi.org/10.5935/0103-5053.20130157.

(40) Thanayutsiri, T.; Patrojanasophon, P.; Opanasopit, P.; Ngawhirunpat, T.; Plianwong, S.; Rojanarata, T. Rapid Synthesis of Chitosan-Capped Gold Nanoparticles for Analytical Application and Facile Recovery of Gold from Laboratory Waste. Carbohydr Polym 2020, 250, 116983. https://doi.org/10.1016/j.carbpol.2020.116983.

(41) Huang, H.; Yang, X. Synthesis of Chitosan-Stabilized Gold Nanoparticles in the Absence/Presence of Tripolyphosphate. Biomacromolecules 2004, 5 (6), 2340–2346. https://doi.org/10.1021/bm0497116.

(42) Sun, L.; Li, J.; Cai, J.; Zhong, L.; Ren, G.; Ma, Q. One Pot Synthesis of Gold Nanoparticles Using Chitosan with Varying Degree of Deacetylation and Molecular Weight. Carbohydr Polym 2017, 178, 105–114. https://doi.org/10.1016/j.carbpol.2017.09.032.

(43) Oliveira, J. P.; Prado, A. R.; Keijok, W. J.; Ribeiro, M. R. N.; Pontes, M. J.; Nogueira, B. V.; Guimarães, M. C. C. A Helpful Method for Controlled Synthesis of Monodisperse Gold Nanoparticles through Response Surface Modeling. Arabian Journal of Chemistry 2020, 13 (1), 216–226. https://doi.org/10.1016/j.arabjc.2017.04.003.

(44) Potara, M.; Maniu, D.; Astilean, S. The Synthesis of Biocompatible and SERS-Active Gold Nanoparticles Using Chitosan. Nanotechnology 2009, 20 (31), 315602. https://doi.org/10.1088/0957-4484/20/31/315602.

(45) Komalam, A.; Muraleegharan, L. G.; Subburaj, S.; Suseela, S.; Babu, A.; George, S. Designed Plasmonic Nanocatalysts for the Reduction of Eosin Y: Absorption and Fluorescence Study. Int Nano Lett 2012, 2 (1), 26. https://doi.org/10.1186/2228-5326-2-26.

(46) Papadakis, G.; Pantazis, A. K.; Fikas, N.; Chatziioannidou, S.; Tsiakalou, V.; Michaelidou, K.; Pogka, V.; Megariti, M.; Vardaki, M.; Giarentis, K.; Heaney, J.; Nastouli, E.; Karamitros, T.; Mentis, A.; Zafiropoulos, A.; Sourvinos, G.; Agelaki, S.; Gizeli, E. Portable Real-Time Colorimetric LAMP-Device for Rapid Quantitative Detection of Nucleic Acids in Crude Samples. Sci Rep 2022, 12 (1), 3775. https://doi.org/10.1038/s41598-022-06632-7.

(47) Zhang, X.; Servos, M. R.; Liu, J. Ultrahigh Nanoparticle Stability against Salt, PH, and Solvent with Retained Surface Accessibility via Depletion Stabilization. J Am Chem Soc 2012, 134 (24), 9910–9913. https://doi.org/10.1021/ja303787e.

(48) Lang, N. J.; Liu, B.; Zhang, X.; Liu, J. Dissecting Colloidal Stabilization Factors in Crowded Polymer Solutions by Forming Self-Assembled Monolayers on Gold Nanoparticles. Langmuir 2013, 29 (20), 6018–6024. https://doi.org/10.1021/la3051093.

(49) Uribe-Alvarez, C.; Lam, Q.; Baldwin, D. A.; Chernoff, J. Low Saliva PH Can Yield False Positives Results in Simple RT-LAMP-Based SARS-CoV-2 Diagnostic Tests. PLoS One 2021, 16 (5), e0250202. https://doi.org/10.1371/journal.pone.0250202.

(50) Pandey, P.; Reddy, Nv.; Rao, V.; Saxena, A.; Chaudhary, C. Estimation of Salivary Flow Rate, PH, Buffer Capacity, Calcium, Total Protein Content and Total Antioxidant Capacity in Relation to Dental Caries Severity, Age and Gender. Contemp Clin Dent 2015, 6 (5), 65. https://doi.org/10.4103/0976-237X.152943.

