## Supporting Information for "Naked-Eye Detection of LAMP-Produced Nucleic Acids in Saliva using Chitosan-capped AuNPs in a Single-Tube Assay"

### Experimental Procedures

#### 1. Reagents and materials

Gold (III) chloride trihydrate ($HAuCl_4$ x $3H_2O$, ACS reagent, >=49% Au basis), chitosan low molecular weight (LMW, 50-190 kDa, 75-85% deacetylated), acetic acid (glacial, ACS reagent, >= 99.7%), water for chromatography (LC-MS grade) LiChrosolv® and phosphate buffer saline (PBS) tablets were purchased from Merck (Darmstadt, Germany). All chemicals were used as received, without any further purification.
The set of six primers (100 μM) for the *Salmonella* invasion gene invA were purchased from Metabion (Germany), where their sequences (5'-3') were as follows:[1]
Inner FIP: *GACGACTGGTACTGATCGATAGTTTTTCAACGTTTCCTGCGG*
Inner BIP: *CCGGTGAAATTATCGCCACACAAAACCCACCGCCAGG*
Outer F3: *GGCGATATTGGTGTTTATGGGG*
Outer B3: *AACGATAAACTGGACCACGG*
Loop F: *GACGAAAGAGCGTGGTAATTAAC*
Loop B: *GGGCAATTCGTTATTGGCGATAG*
The set of six primers (100 μM) for the SARS-CoV-2 detection targeting the N gene were purchased from Eurofins Genomics (Germany), where their sequences were as follows:[2]
Inner FIP: *TGCGGCCAATGTTTGTAATCAGCCAAGGAAATTTTGGGGC*
Inner BIP: *CGCATTGGCATGGAAGTCACTTTGATGGCACCTGTGTAG*
Outer F3: *AACACAAGCTTTCGGCAG*
Outer B3: *GAAATTTGGATCTTTGTCATCC*
Loop F: *TTCCTTGTCTGATTAGTTC*
Loop B: *ACCTTCGGGAACGTGGTT*
Warmstart® Multi-Purpose LAMP/RT-LAMP 2X Master Mix, WarmStart® Colorimetric LAMP 2X Master Mix (DNA & RNA) and LAMP fluorescent dye (readable in the SYBR®/FAM channel of real-time fluorimeters) were purchased from New England BioLabs. Normal saliva (pooled human donors) was purchased from Lee Biosolutions, USA. Mineral oil (BioReagent, for molecular biology) was purchased from Merck. Synthetic SARS-CoV-2 RNA was purchased from BIORAD (SARS-CoV-2 Standard #COV019 and SARS-CoV-2 Negative #COV000).

### 2. Chit-AuNPs synthesis and purification

Chit-AuNPs were synthesized utilizing a chemical reduction process, with chitosan acting as both a reducing and stabilizing agent.[3] Volumes of 15 mL of 0.15%, 0.25% and 0.35% (w/v) LMW chitosan solutions (in 1% (v/v) aqueous acetic acid) were heated to a temperature of 45 °C and stirred for 3 h. Afterwards, the temperature was raised to 75 °C, and 0.15 mL of an aqueous $HAuCl_4$ solution (100 mM) was added to each of the solutions while under stirring. After 2h of vigorous stirring under heating, the solutions were moved away from the heating plate and remained under stirring until room temperature ($T_R$). The solutions were then stored at 4 °C until further use. In all cases, different shades of red color were obtained, indicating the formation of Chit-AuNPs in different sizes/concentrations. The color change from light yellow to red started in the solutions with the higher Chit concentrations first, which means that the reduction process was faster.

Centrifugation was carried out using an Eppendorf 5417R Refrigerated Centrifuge. The 0.15% (w/v) Chit-AuNPs were centrifuged at appropriate conditions (volumes of 0.5 mL at 2000 rcf for 30 min, at 4 °C), in order to remove the free Chit from the solution. Afterwards, redispersion of the pellet was performed in either (a) 1% (v/v) aqueous acetic acid; (b) ultrapure water; or (c) 1% (v/v) aqueous acetic acid containing 0.15% (w/v) Chit. The amount of excess Chit chosen for the redispersion is the same one used for the synthesis, so that it should naturally be higher than the free dissolved non-reacted Chit in the originally synthesized one. This purification step was carried out in order to study the effect of free Chit, as well as the pH of the final LAMP mixtures, after the addition of the Chit-AuNPs by spin-down.

### 3. Chit-AuNPs characterization

A variety of instrumental analytical methods were used to characterize the synthesized Chit-AuNPs. Initially, attenuated total reflectance/Fourier-transform infrared spectroscopy (ATR-FTIR) was used to semi-quantitatively measure the observable IR spectrum of the neat Chit and Chit-AuNPs by evaluating the transmittance over a spectral region of 4000 to 400 $cm^{-1}$. A VERTEX 70v FT-IR Spectrometer (Bruker), equipped with a A225/Q Platinum ATR unit with single reflection diamond crystal was used. To achieve a suitable signal quality, all spectra were collected at a resolution of 4 $cm^{-1}$ by collecting 50 scans. Specimens were prepared by drop coating the studied solutions onto chemically cleaned glass substrates, and drying them at 60 °C for 30 min. The Chit-AuNPs formation was confirmed by observation of the surface plasmon resonance (SPR) band using UV-Visible absorption spectroscopy (Nanodrop ND-1000), by using 2 μL of each sample. The Chit-AuNPs size and shape were studied by field emission gun - scanning electron microscopy (FEG-SEM, JSM-IT700HR, Jeol Ltd., Tokyo, Japan). Specimens for SEM measurements were prepared by drop coating diluted the colloidal solutions onto chemically cleaned silica substrates, and left to dry at ambient conditions. Image analysis of the SEM micrographs was performed using ImageJ software in order to calculate the average diameters of the synthesized Chit-AuNPs, where 100 NPs were measured from each sample. Finally, the z-average (intensity weighted mean hydrodynamic diameter ($D_h$)), polydispersity index (PDI) and mean z-potential values of the studied solutions were determined by dynamic light scattering (DLS) and zeta potential (ZP) measurements, using a Malvern Zetasizer Nano ZS90 (Malvern Instruments Ltd, UK) after diluting them with ultrapure water, while measurements were performed in triplicate.

### 4. Synthesized Chit-AuNPs molar concentration calculation

The mean core size of the three Chit-AuNPs solutions was calculated by analyzing the SEM micrographs using the ImageJ software, as previously mentioned. The mean Chit-AuNPs diameter (D) calculated were: 38.4, 13.8 and 10.7 nm for Chit-AuNPs synthesized using 0.15%, 0.25% and 0.35% (w/v) Chit, respectively. For the Chit-AuNPs concentration calculation, three assumptions are made: i) $HAuCl_4$ is entirely converted into AuNPs, ii) density (ρ) of AuNPs is equal to density of bulk Au (19.3 g/cm$^3$) and iii) AuNPs are spherical in shape with a uniform FCC crystal structure.[4] The average number of gold atoms (N) per nanoparticle can be expressed as:

$$N = \frac{\pi \rho N_A D^3}{6M}$$

where M= 197 g/mol the molar mass of Au and $N_A$: Avogadro's number, while the concentration (C) of the nanoparticles inside the solution as:

$$C = \frac{moles\ from\ reaction}{NV}$$

where V: total reaction volume (in liters).

The molar concentrations (C) were calculated as: 0.57, 12.3 and 26.4 nM for Chit-AuNPs synthesized using 0.15%, 0.25% and 0.35% (w/v) Chit, respectively.

### 5.Colorimetric LAMP assays preparation and evaluation

Evaluation experiments were performed with an attenuated strain of *Salmonella* enterica serovar Typhimurium as target. *Salmonella* was grown overnight in Luria-Bertani (LB) medium and the cultures were subsequently measured spectrophotometrically ($OD_{600}$), to an $OD_{600}$ of 0.23 corresponding to a cell concentration of 3x10$^8$ cfu/mL. Cell lysis (heating at 95 °C for 10 min) was performed in the *Salmonella* cells stock solution before further use, while serial dilutions using PBS were carried out to reach the final required concentrations. PBS solution (0.01 mol/L, pH 7.4) was prepared by dissolving a PBS tablet in 1 L of ultrapure water. For the purified samples, the LAMP reagent mixes in a total volume of 25 μL contained 12.5 μL Warmstart, 2.5 μL of primer mix (containing 18 μM FIP and BIP, 2 μM F3 and B3 and 6 μM Loop-F and Loop-B), 9 μL of nuclease-free water and 1 μL of the target at appropriate dilutions (as negative control, 1 μL of PBS solution was used). For the crude saliva samples, the LAMP reagent mix was similar, but 2.5 and 5 μL of lysed saliva (95 °C for 10 min) replaced a corresponding amount of nuclease-free water in the final reaction's volume. Cross-study in RT-LAMP assay using SARS-CoV-2 as target was carried out with the corresponding reagents amounts similar to the LAMP-*Salmonella* crude saliva samples assays. In this case, the dilutions from the target's starting stock to different SARS-CoV-2 copies were performed in nuclease-free water (as negative control, 1 μL of neat nuclease-free water was used). Volumes of 7.5 μL of Chit-AuNPs solutions were immobilized inside the tube's lid by surface tension, in order to not interfere with the LAMP reagents, as well as to avoid aerosol contamination. After the LAMP reaction was complete (at T=63 °C for t=30 min in *Salmonella* assay and T=65 °C for 30 min in SARS-CoV-2 assay), a brief spin-down (~10 s) lead to the mixture of the two different solutions, and the result was ready to be read colorimetrically by naked eye. For the Chit-AuNPs based assays, heating of the 0.2 mL PCR tubes (Sarstedt) was performed using a FastGene Ultra Cycler Gradient (Nippon GENETICS Europe), where no heating on top of the tubes was applied as it could evaporate the immobilized Chit-AuNPs solutions. Real time quantitative colorimetric LAMP (qcLAMP) using phenol red (pH indicator) was performed using Pebble (Biopix-T, Gr),[5] where an addition of 15 μL mineral oil was added over the LAMP mix, in order to avoid solvent evaporation owing to tubes' heating. Finally, Coyote Mini8 Plus Real Time PCR Cycler (Coyote Bioscience Co., Ltd., Beijing, China) was used for the real time fluorescent LAMP assays.

### Supporting Figures

#### 1. LAMP reaction in the presence of Chit-AuNPs

Addition of neat Chit or Chit-AuNPs directly inside the LAMP mix completely inhibits the amplification reaction (S1a), owing to their electrostatic interactions with LAMP reagents (final solution pH~4.5). In these reactions, 5 μL of nuclease-free water were replaced by either 0.15% (w/v) neat Chit or Chit-AuNPs, and 0.5 μL of nuclease-free water by 0.5 μL of LAMP fluorescent dye. Picture (S1b) shows the tubes after the LAMP reaction. In the normal fluorescence assay, the negative sample remains transparent while the positive (100 cfu/reaction *Salmonella*) is blurrier owing to the increase of turbidity because of LAMP's $Mg_2P_2O_7$ by-product.[6] When incorporating neat Chit inside the solutions, both negative and positive samples appear blurry due to the presence of free Chit. In the Chit-AuNPs case, a visible pellet is formed possibly due to the Chit-AuNPs heat-induced aggregation inside the LAMP mix, as Chit-AuNPs obtain enough energy (63 °C heating) to cross the free Chit's depletion barrier.[7]

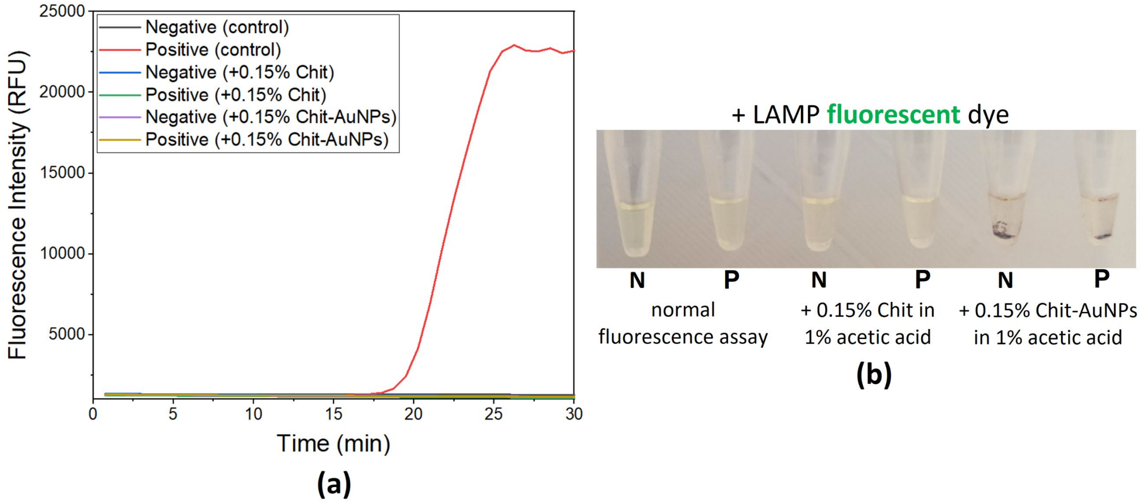


***Figure S1:*** *a) Real time fluorescence LAMP using the fluorescent dye. Addition of neat Chit or Chit-AuNPs completely inhibits the LAMP reaction. b) Photograph of the tubes after LAMP reaction. No apparent colorimetric difference can be observed between the negative and positive samples when neat Chit or Chit-AuNPs are included, due to the inhibition of the amplification reaction. Note: The N down the photograph of the 0.2 mL tubes corresponds to negative samples, while the P to positives with 100 cfu/reaction (Salmonella).*

#### 2. Purification of Chit-AuNPs via centrifugation

The Chit-AuNPs synthesized using 0.15% (w/v) Chit were centrifuged, in order to purify the solution by removing the free dissolved/non-reacted Chit. After redispersion in the different chosen media, no color change was observed, indicating no aggregation during the purification step. This is also confirmed via the UV-Vis spectra (S2a), where no red shifting in the SPR $\lambda_{max}$ was observed. Furthermore, measurements of the $D_h$ and z-potential (S2b) of the resulting solutions revealed a significant increase in the case of excess Chit addition in both parameters (159.7 nm mean $D_h$ and +68.4 mV z-potential) compared to the originally synthesized solution (59.6 nm mean $D_h$ and +45.7 mV z-potential), possibly reflecting free Chit's dominating presence inside the solution. The solutions without free Chit (redispersion in 1% (v/v) aqueous acetic acid or ultrapure water) didn't present significant differences in the z-potential values when compared to the originally synthesized solution, while the $D_h$ showed a decrease of about 16 nm in both cases.

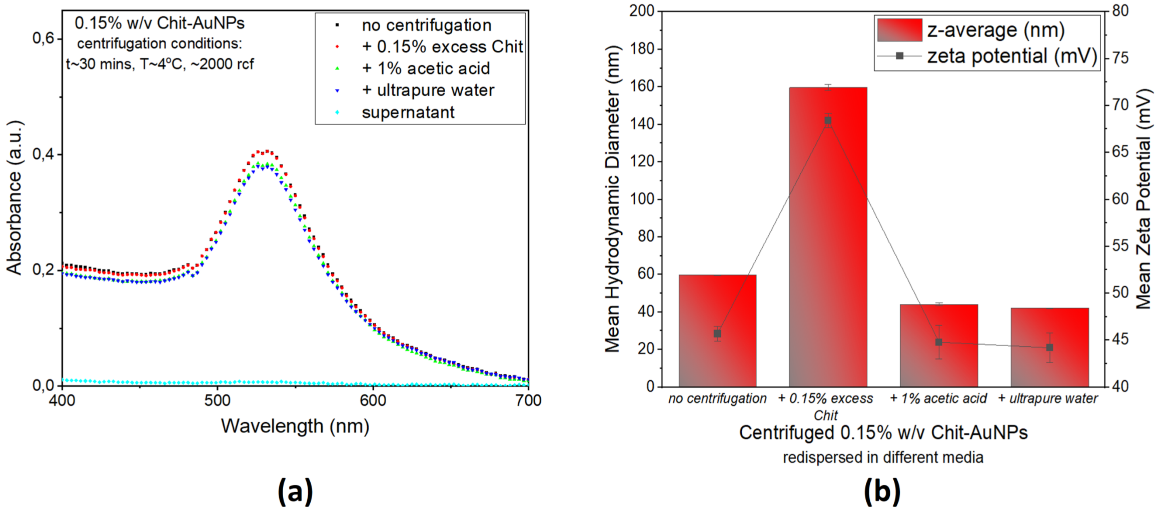


***Figure S2:*** *a) UV-Vis spectra, b) DLS and ZP measurements, of the 0.15% (w/v) Chit-AuNPs, after centrifugation and redispersion in different solvents (0.15% (w/v) Chit in 1% (v/v) aqueous acetic acid, 1% (v/v) aqueous acetic acid, ultrapure water).*

### 3. Effect of time and pH on the stability of the final solutions

An ageing test in assays using 20% saliva revealed that when using free Chit, the negatives also tend to aggregate after some time (~4 days), possibly due to the differently charged populations inside the solution. At the same time, the aggregation in the positive samples happened significantly faster (immediately after the spin-down/mixing of the Chit-AuNPs and LAMP solutions), therefore there was no problem differentiating the positive samples from the negative ones. In this assay, the pellet formed is initially a Chit-DNA electrostatic complex,[8] which subsequently is further enriched by Chit-AuNPs, and hence the pellet appearance inside the tube. In the assays without free Chit, the positive samples remain stable over time, owing to DNA-induced electrosteric stabilization of Chit-AuNPs inside the LAMP mix.[7] In this assay, the pellet formed in the negative samples is due to Chit-AuNPs aggregation induced by the different LAMP reagents, as there is not enough DNA present in the solution to efficiently stabilize them by electrostatic conjugation (S3a). In the case of removal of free Chit and redispersion of 0.15% (w/v) Chit-AuNPs in ultrapure water instead of 1% (v/v) aqueous acetic acid, aggregation in both negative and positive samples was observed. This indicates that when the pH of the final solution (Chit-AuNPs + LAMP) is close to or higher than Chit's pKa of ~6.5,[9] the electrostatic interactions between the Chit-AuNPs and the amplified DNA are not favored due to the negligible Chit-AuNPs charge, and the Chit-AuNPs also tend to aggregate in the positive samples owing to the dominating attractive Van der Waals forces inside the LAMP mix (S3b).

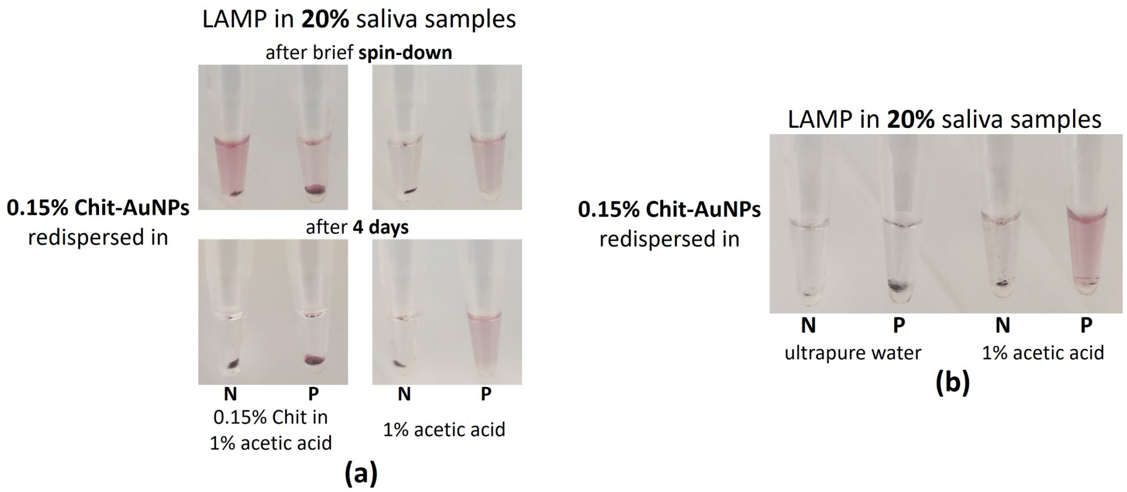


***Figure S3:*** *a) Effect of time on the stability of LAMP reaction-Chit-AuNPs mix after amplification, in the presence of 20% saliva samples. In the assay using free Chit, the stable negative is gradually destabilized over time, while in the assay without free Chit, the stable positive maintains its stability over time. b) Effect of pH on the redispersion medium of centrifuged 0.15% (w/v) Chit-AuNPs. An acidic pH is preferable instead of a neural for a visible colorimetric difference between negative and positive samples, in order for electrostatic interactions between Chit-AuNPs and DNA to occur. Note: The N down the photographs of the 0.2 mL tubes corresponds to negative samples, while the P to positives with 100 cfu/reaction (Salmonella).*

### 4. Zeta-potential and absorbance of the final stable solutions

In the assay using Chit-AuNPs with free Chit, the stable negative sample presents a positive z-potential value (~ +10.7 mV), meaning that the supernatant's stability is owing to the dominating presence of free Chit in the solution which hasn't been completely saturated. In the assay using Chit-AuNPs only, the stable positive sample presents a negative z-potential value (~ -7.1 mV), indicative of the efficient electrostatic conjugation of positively charged Chit-AuNPs with negatively charged amplified DNA (S4a). This conjugation is further confirmed by the observed red-shifting in the UV-Vis spectra, where the stable positive solution presented a redshift of the SPR $\lambda_{max}$ to 541 nm and a drop in the absorbance intensity, compared to the stable negative sample that presented a $\lambda_{max}$ of 532 nm (S4b).

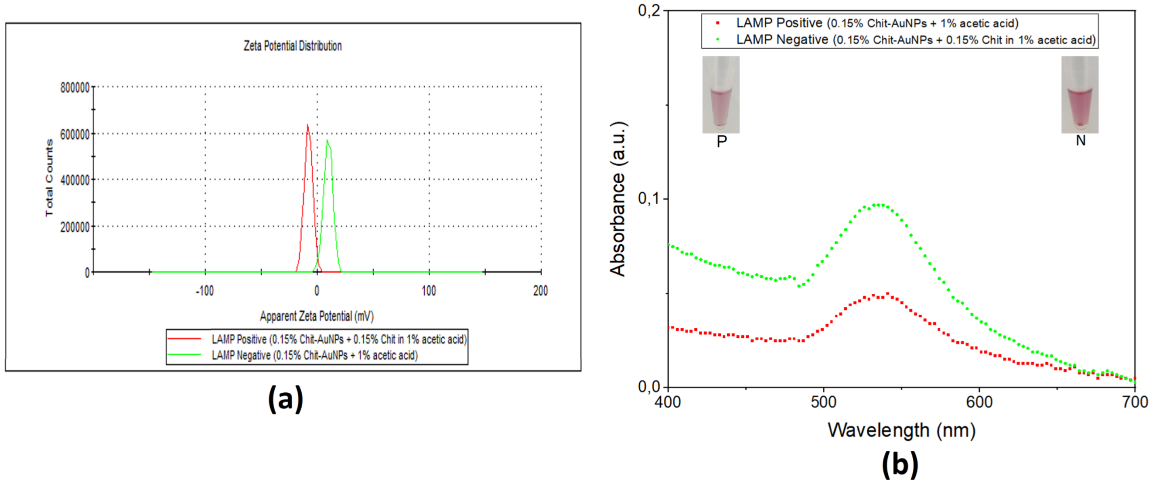


***Figure S4:*** *a) Zeta potential distribution and b) UV-Vis absorbance spectra, of a stable negative (free Chit/Chit-AuNPs + LAMP) and a stable positive (Chit-AuNPs + LAMP) sample. The stable negative presented a positive z-potential (~ +10.7 mV) and an SPR $\lambda_{max}$ at 532 nm, while the stable positive presented a negative z-potential (~ -7.1 mV) and an SPR $\lambda_{max}$ at 541 nm, indicative of the efficient conjugation of the amplified DNA to the Chit-AuNPs surface in the latter case.*

### References


(1) Hara-Kudo, Y.; Yoshino, M.; Kojima, T.; Ikedo, M. Loop-Mediated Isothermal Amplification for the Rapid Detection of Salmonella. *FEMS Microbiol Lett* **2005**, *253* (1), 155–161. https://doi.org/10.1016/j.femsle.2005.09.032.

(2) Broughton, J. P.; Deng, X.; Yu, G.; Fasching, C. L.; Servellita, V.; Singh, J.; Miao, X.; Streithorst, J. A.; Granados, A.; Sotomayor-Gonzalez, A.; Zorn, K.; Gopez, A.; Hsu, E.; Gu, W.; Miller, S.; Pan, C.-Y.; Guevara, H.; Wadford, D. A.; Chen, J. S.; Chiu, C. Y. CRISPR–Cas12-Based Detection of SARS-CoV-2. *Nat Biotechnol* **2020**, *38* (7), 870–874. https://doi.org/10.1038/s41587-020-0513-4.

(3) Lakshmi Narayanan, R.; Sivakumar, M. Preparation and Characterization of Gold Nanoparticles in Chitosan Suspension by One-Pot Chemical Reduction Method. *Nano Hybrids* **2014**, *6*, 47–57. https://doi.org/10.4028/www.scientific.net/NH.6.47.

(4) Khashayar, P.; Amoabediny, G.; Larijani, B.; Hosseini, M.; Vanfleteren, J. Fabrication and Verification of Conjugated AuNP-Antibody Nanoprobe for Sensitivity Improvement in Electrochemical Biosensors. *Sci Rep* **2017**, *7* (1), 16070. https://doi.org/10.1038/s41598-017-12677-w.

(5) Papadakis, G.; Pantazis, A. K.; Fikas, N.; Chatziioannidou, S.; Tsiakalou, V.; Michaelidou, K.; Pogka, V.; Megariti, M.; Vardaki, M.; Giarentis, K.; Heaney, J.; Nastouli, E.; Karamitros, T.; Mentis, A.; Zafiropoulos, A.; Sourvinos, G.; Agelaki, S.; Gizeli, E. Portable Real-Time Colorimetric LAMP-Device for Rapid Quantitative Detection of Nucleic Acids in Crude Samples. *Sci Rep* **2022**, *12* (1), 3775. https://doi.org/10.1038/s41598-022-06632-7.

(6) Mori, Y.; Kitao, M.; Tomita, N.; Notomi, T. Real-Time Turbidimetry of LAMP Reaction for Quantifying Template DNA. *J Biochem Biophys Methods* **2004**, *59* (2), 145–157. https://doi.org/10.1016/j.jbbm.2003.12.005.

(7) Shin, J.; Zhang, X.; Liu, J. DNA-Functionalized Gold Nanoparticles in Macromolecularly Crowded Polymer Solutions. *J Phys Chem B* **2012**, *116* (45), 13396–13402. https://doi.org/10.1021/jp310662m.

(8) Amaduzzi, F.; Bomboi, F.; Bonincontro, A.; Bordi, F.; Casciardi, S.; Chronopoulou, L.; Diociaiuti, M.; Mura, F.; Palocci, C.; Sennato, S. Chitosan-DNA Complexes: Charge Inversion and DNA Condensation. *Colloids Surf B Biointerfaces* **2014**, *114*, 1–10. https://doi.org/10.1016/j.colsurfb.2013.09.029.

(9) de Oliveira, A. C.; Sabino, R. M.; Souza, P. R.; Muniz, E. C.; Popat, K. C.; Kipper, M. J.; Zola, R. S.; Martins, A. F. Chitosan/Gellan Gum Ratio Content into Blends Modulates the Scaffolding Capacity of Hydrogels on Bone Mesenchymal Stem Cells. *Materials Science and Engineering: C* **2020**, *106*, 110258. https://doi.org/10.1016/j.msec.2019.110258.
